# Implementation of a Technology-driven Antimicrobial Stewardship Program Steered by Clinicians to Improve Antimicrobial Prescribing: protocol of a multi-centre stepped wedge trial

**DOI:** 10.1101/2025.03.27.25324784

**Authors:** Archana Siddaiah, Carol D’Silva, Bharat Kalidindi, N Thenmozhi, Manjulika Vaz, Savitha Nagaraj

## Abstract

**Background:** The World Health Organization (WHO), Centre for disease Control and Prevention (CDC) gives broad guidance on how to establish, implement and evaluate AMSP. However, specific action plans for effective AMSP especially in LMICs is needed, the action plan in India is not uniformly implemented across hospitals because of prevailing issues specific to diverse hospital settings. These include-non-availability of all classes of antimicrobial agents (AMA) in the hospital, lack of in-house antibiotic policy which may lead to irrational prescription and lack of skilled manpower such as clinical pharmacists who are the pillars to prescription audits. Crucial to all this is how clinical teams are constantly engaged in informed AMA prescribing. As per the NAP-AMR strategy, hospitals have been trying to implement AMSP inspite of resource constraints. Ensuring only right drugs are used at the right time, is challenging because, engaging clinical teams has been an important bottleneck.

**Objectives:** To explore the key promoters, constraints, and operational feasibility of an integrated m-health intervention program on antimicrobial consumption and AMSP in five tertiary hospitals in south India; To evaluate the effectiveness of an integrated m-health intervention program on antimicrobial consumption and AMSP in five tertiary hospitals in south India; To assess the impact of an integrated AMSP m-health intervention on the incidence of multidrug-resistant organisms in five tertiary hospitals in south India

**Methodology:** Study will be conducted in four tertiary hospitals across south India. Pre-intervention-A baseline data collection will be done before the delivery of the intervention. Intervention: This includes capacity building of clinicians on AMSP and provision of mobile application for them to use during patient care. Implementation of intervention-A stepped wedge trail will be conducted in the selected units various departments included in the study. This will be done over 24 months. All units receiving the intervention will be followed up for the next eight months periodically.

**Outcomes:** Outcome indicators such as consumption of antimicrobial agents, incidence of multi drug resistance organisms and healthcare associated infections will be captured during the follow ups.

## INTRODUCTION

Clinicians play a significant role in consumption of antimicrobials by routine prescriptions. Appropriate prescription of antimicrobials is crucial for positive clinical outcomes in patients. However, the emergence of antimicrobial resistance (AMR) has become a public health threat.(1) Infections due to resistant pathogens are responsible for a high healthcare burden and are estimated to cause over 7,00,000 deaths annually worldwide– a rate which is projected to rise to 10 million by 2050.(2,3) AMR depends on how antimicrobials are prescribed and consumed. 78% of antibiotics in low- and middle-income countries being self-medicated and unregulated. Also, it is increasingly harder to monitor antibiotic use in these countries. (4–6) India is one of the countries among LMICs that is the largest consumer of antibiotics.(7,8) According to estimates, more than 50% of hospital antibiotic use is inappropriate in India.(9) This is a compelling indication for an urgent augmented education and awareness program on AMR among clinicians.

The World Health Organization (WHO) defines anti-microbial stewardship (AMS) as a coherent set of integrated actions which promote the responsible and appropriate use of antimicrobials to help improve patient outcomes across the continuum of care. It has identified five pillars of AMS programme. One of these includes strengthening the health worker capacity through the provision of tailored education and training packages.(10) Taking cognizance of the AMR situation, the Indian Ministry of Health and Family Welfare launched the National Action Plan (NAP) for AMR in 2017.(11) Efforts are ongoing by the National Medical Commission in implementing a competency based medical curriculum on AMS, mainly aimed at the medical students, who will be the future prescribers.(12) However, given the enormous threat that AMR poses to public health, we cannot risk relying on interventions having long term outcomes. This is a time to take swift and immediate corrective actions.

Currently, a lack of knowledge on AMR among healthcare practitioners has been identified as one of the barriers that impacted the antimicrobial prescriptions.(13) This also included a gap in the knowledge and perception of optimal antibiotic prescription practices.(14) Studies have looked at using technological solutions such as digital support algorithms for effective antimicrobial prescribing that can overcome such barriers.(15) A study from United Kingdom used health information technology for timely review of antimicrobial therapies and stop unnecessary antibiotics. A systematic review that investigated stewardship smartphone applications used by physicians found studies mostly from developed countries, and none of these studies were randomized controlled trials.(16) Majority of the included studies looked at the process indicators, adherence to guidelines, user experience, and antimicrobial consumption. A metanalysis also found a positive association with the clinical decision support system and appropriateness of antibiotic therapy and had a positive effect on adherence to guidelines. (17) Smartphone apps were found to be an acceptable format to deliver trustworthy guidance on antimicrobial prescribing in an accessible format at the bedside and they promoted adherence to hospital guidelines.(16,18,19)

There are many gaps in effective implementation and monitoring of the WHO guidelines on AMS especially in LMICs. As per this NAP, hospitals across India have been trying to implement AMSP in spite of resource constraints. Also, these constraints vary across different settings. Some of which include-absence of in-house antibiotic policy non-availability of all classes of antimicrobials in the hospital formulary leading to non-adherence to the hospital policy, lack of antibiogram, lack of skilled manpower such as clinical pharmacists who are an integral component of multi-disciplinary team to strengthen AMS program.(20) Crucial to all this is how clinical teams are constantly engaged in informed antimicrobial prescribing. Engaging clinical teams has been an important bottleneck.(21,22)

### Rationale

The AMS interventions can be broadly classified into interventions prior to/at the time of prescription and interventions after prescription.(23) The clinician education, institutional guidelines, and use of antibiograms comes under the former category. The current proposal aims to evaluate a technological intervention prior to/at the time of prescription using the above listed components through the clinician-patient encounter framework. To the best of our knowledge, the current proposal is one of the first to evaluate the effectiveness of a m-health intervention in improving the AMS program in our context using a stepped wedge cluster randomized controlled trial (SWT). The study adopted a (SWT) design due to its strong potential to minimize contamination, particularly in the context of a behavioral intervention. Also, because we wanted to test the intervention in the routine context as opposed to controlled settings, we feel such pragmatic design is most suitable. Therefore, we hypothesize that an integrated m-health AMS intervention in the form of a smartphone app containing institutional antibiotic policy and antibiogram complemented by capacity building of clinicians on AMS program will promote rationale prescription of antimicrobials and in turn improve patient outcomes. The benefit of such an integrated app is that it will give clinicians immediate access to the hospital policy and antibiograms. The study will also shed light on the feasibility and sustainability of implementing such intervention and strengthen AMS within the context of both public and private hospitals. Thus, learnings from this study will help in understanding the operational feasibility in scaling up AMS interventions across India.

### Objectives

1. To explore the key promoters, constraints, and operational feasibility of an integrated m-health intervention program on antimicrobial consumption and AMSP in four tertiary hospitals in south India.
2. To evaluate the effectiveness of an integrated m-health intervention program on antimicrobial consumption and AMSP in four tertiary hospitals in south India.
3. To assess the long-term impact of an integrated AMSP m-health intervention on the incidence of multidrug-resistant organisms in four tertiary hospitals in south India.

### Study setting

Four tertiary academic medical institution in south India will be included having a mix of public and private set up. Four study hospitals are located in the state of Karnataka and two are in the state of Tamil Nadu.

For the objective 1 we will do a series of qualitative interviews with concerned stakeholders, situational analysis of the existing AMS program in the study hospitals. Also, assessment of clinicians’ needs to come up with a m-health intervention that can facilitate AMS program will also be done in this phase. Stakeholders for qualitative interviews include hospital administrators, managers, clinicians working in infectious diseases, critical care departments, hospital antibiotic committee members, infection control committee members, clinical pharmacists, microbiologists, patient groups, and nursing personnel. Purposive sampling will be done to select participants for qualitative discussions. Those who are working in their respective roles for atleast an year will be eligible to participate in the interviews.

## METHODS (objective 2)

### Trial design

This study will be employ a multi-centre sequential SWT. Each study hospital will serve as a cluster with two fixed departments. Within each department there will be a number of units that will be included in the SWT. A total of four clusters (hospitals) having a total of 38 units will be part of the SWT. Four clusters will be allotted to each of the five sequences over a period of one year. The time duration between each step i.e, cross over from control to the intervention arm will be three months. Study participants will be drawn from each of the selected units. Study participants assessed in different time periods will be the same. However, patient data from each units will be arising from different group of patients at different time periods.

### Eligibility criteria for clusters and participants

#### Settings and locations where the data were collected

The SWT will be conducted in four tertiary care teaching hospitals located in two different states of India. Two of these hospitals are located in Bengaluru, Karnataka and one each in Chennai and Trichy in the state of Tamil Nadu. Among these hospitals, two belong to the public sector.

We will select participants from the clinical departments that will be selected based on the past antimicrobial consumption data. Departments with the highest antimicrobial use (two departments from each study hospital) will be selected for the SWT. The selected departments that will be included in the SWT are given in **Table 2**.

**Table 1:**
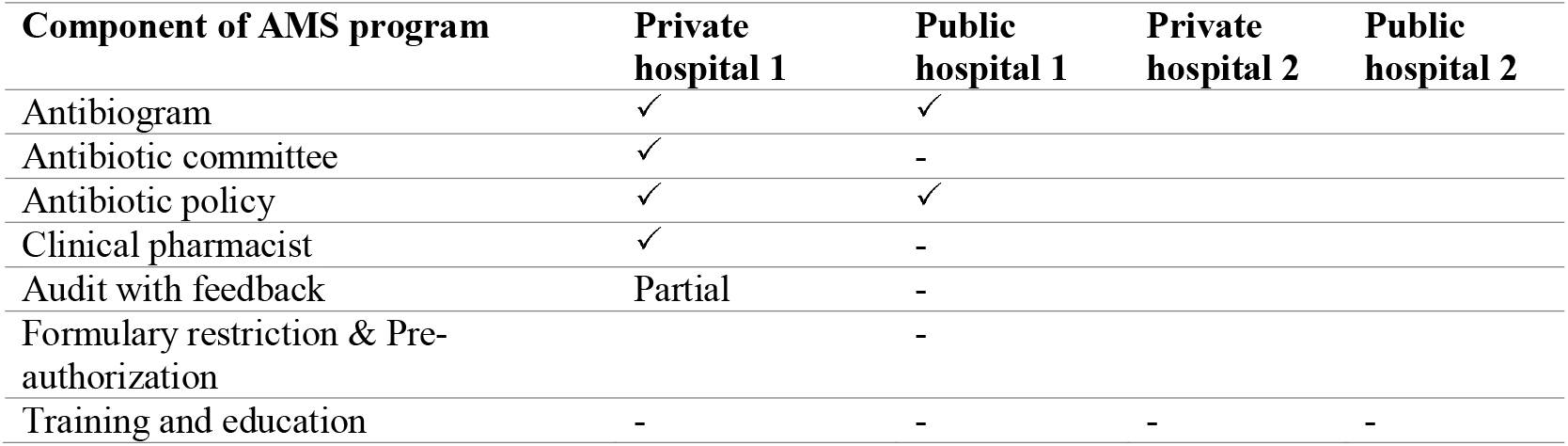
Status of AMS program across the study hospitals.

| Component of AMS program | Private hospital 1 | Public hospital 1 | Private hospital 2 | Public hospital 2 |
| --- | --- | --- | --- | --- |
| Antibiogram | ✓ | ✓ |  |  |
| Antibiotic committee | ✓ | - |  |  |
| Antibiotic policy | ✓ | ✓ |  |  |
| Clinical pharmacist | ✓ | - |  |  |
| Audit with feedback | Partial | - |  |  |
| Formulary restriction & Pre-authorization |  | - |  |  |
| Training and education | - | - | - | - |

**Table 2:** Selected departments under each study hospital.

| Clusters | Departments | No. of Units |
| --- | --- | --- |
| Hospital 1 | Medicine | 3 |
|  | Critical care | 2 |
| Hospital 2 | Medicine | 7 |
|  | Surgery | 6 |
| Hospital 3 | Medicine | 6 |
|  | Critical care | 1 |
| Hospital 4 | Medicine | 4 |
|  | Surgery | 6 |

Eligibility criteria for each department will be the antimicrobial consumption pattern as well as the number of clinicians who can prescribe antibiotics. Each department consists of different units with senior consultants, senior residents, junior residents, and rotating medical interns. The number of senior residents is not fixed. All of them will be involved in antimicrobial prescribing. However, the jun or clinicians such as interns and junior residents will take orders from their senior consultants for some of these prescriptions. For the SWT all clinicians working in the selected departments and the units for more than a year will be eligible to participate. Along with this, those who will continue to work in the study hospital for atleast another year will be included in the study.

**Fig 1:**
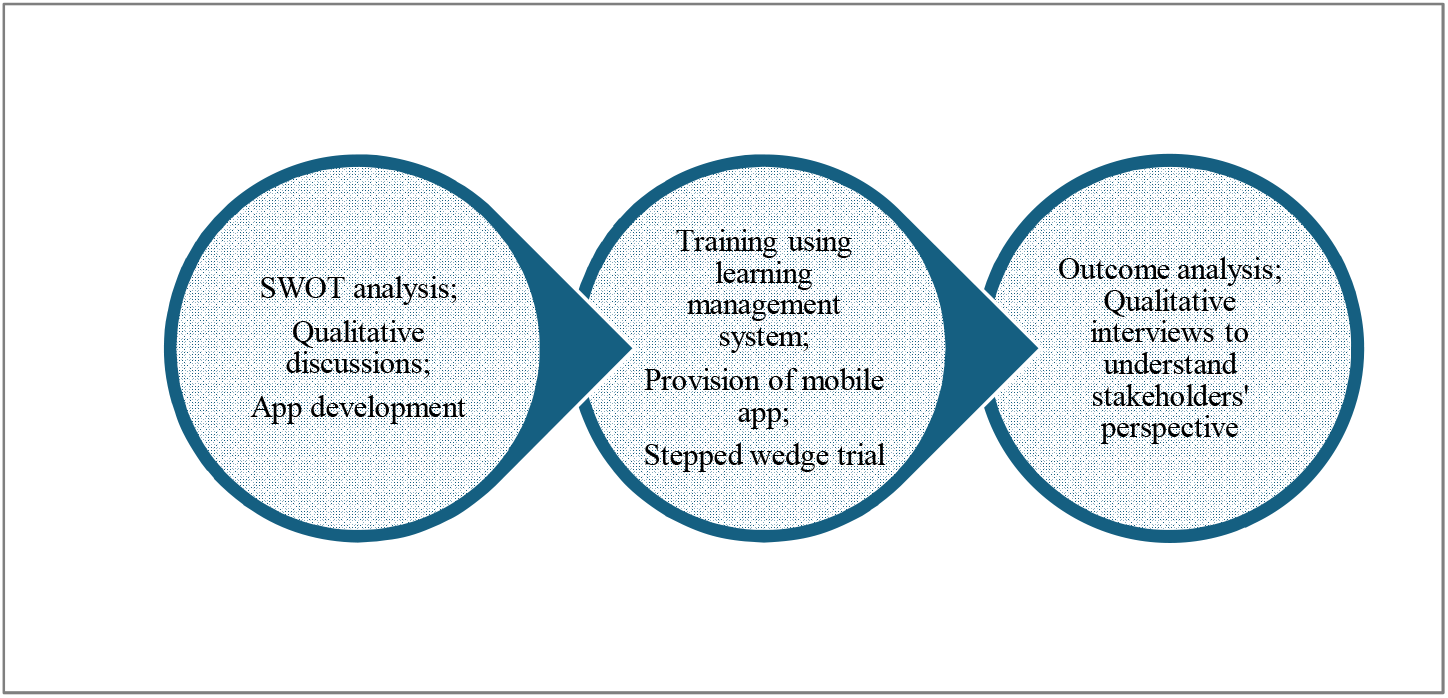
Schemtaic diagram of the study

#### Interventions

Intervention will be delivered at the cluster level. Participants in each cluster will receive the intervention as per the sequence generated. Intervention includes three components. These include training module for clinicians, capacity building of clinicians, and providing m-health mobile application to clinicians which will have the institution-specific antibiotic policy, antibiogram, e-learn module based on the national guidance on antimicrobial use the app and.

### Development of intervention

a. *AMSP training module*-a brief interactive training guide will be prepared by the research team along with clinicians. Contents will be prepared based on the WHO and NAP-AMR guidelines.(5,6,11,12) Materials from the Tackling AMR from the Fleming fund, e-book AMSP From principles to Practice by the British Antimicrobial and Chemotherapy society will be used as reference materials.(13,14)
b. *AMSP mobile application*: The Medical Informatics team from the PI study site will undertake the need assessment from all the stakeholders including the users of the application. All the findings/requirements from the need assessment exercise will be drafted in a Functional Requirements Specification (FRS) Document which will be the blueprint for the application development. This FRS document will be finalized with the research team and will be shared to the application developers to initiate the development process. The Development team will develop the modules and incorporate the features during the App development process. Multimedia content required will be plugged into the app. The development team will also provide a testing app for identification of bugs and user acceptance testing which will be carried out by the medical informatics team. The process will be documented, and any errors, changes and updates will be incorporated during this process. The updated and live version of the m-health application will be released on the google play store and Apple app store which will be utilised by the study team and other users. The app and the database (containing the study data) will be hosted on a secure cloud server, and this will be managed with appropriate guard rails to ensure data security and privacy. Only the Principal Investigator (PI) will have access to the data on the cloud server and may assign limited rights to other investigators based on their role and needs for the study. During the period of the study, the app development and medical informatics team will be providing support to ensure that the app is fully functional, and any issue with it is resolved. Soon after the training, AMSP mobile app will be launched, and clinicians from the intervention units will be asked to download the app and self-register. The registration requests will be approved by the site investigators thereby limiting the access of the app only to the study participants. They will be encouraged to use the app during patient care.
c. *E-learn module in the application*: The research team will develop the video tutorials for the e-learn module. The contents will be uploaded to the app regularly. The research team will track the user progress on the learning content. **Box 1 and 2**.

### Randomization

#### Allocation and Sequence generation

In SWT, the study clusters cross over to the intervention condition at predetermined time points in a sequential, staggered fashion until all groups receive the intervention The selection of department units will be done as per the eligibility criteria. Randomization will be done at the hospital level. Each hospital will be considered as a cluster and within each hospital there will be group of units arising from the selected departments. Allocation sequence will be independently generated by the study biostatistician using block randomization method/ or random ordering of the allocation sequence will be determined from computer-generated uniform random numbers. All hospitals start the trail in the control condition. The generated sequence will determine the time when each study hospital crosses to the intervention arm until all units receive the intervention. As the hospital goes into the intervention, all clinicians working in all the units will receive training and mobile app as described earlier. All clusters will be followed up for eight months.

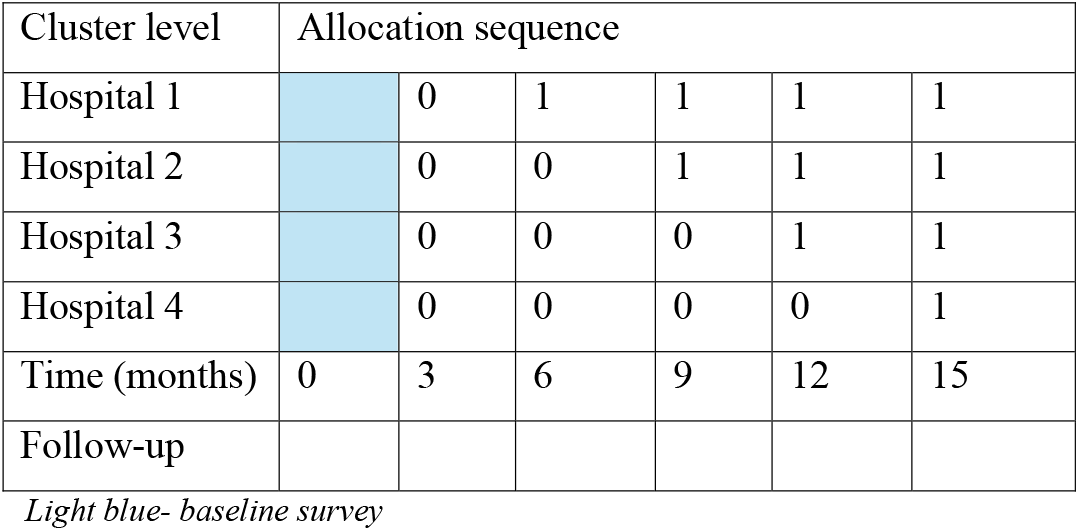
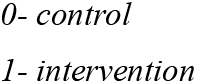

#### Allocation concealment mechanism

The allocation sequence will be known to the study coordinating site. The study participants and the clusters will be blinded to the allocation sequence. Those not receiving yet receiving the intervention will not be aware of the time at which they will have the intervention implemented. Each hospital will come to know about the sequence only at the time of crossing over.

#### Implementation

Randomization schedule will be generated by the study statistician. Enrollment of clusters will be done by the site investigator. In each hospital, departments having the highest antibiotic consumption will be enumerated. Out of this 2-3 will be selected based on the consent of the department and their willingness to participate in the SWT. These clusters will be assigned to the generated sequence by the statistician along with the PI.

#### Recruitment

The recruitment process will happen simultaneously at all hospitals. For this study, all clinicians working in the selected departments will be recruited. A complete enumeration of all clinicians will be done with the help of the head of the departments. Based on the inclusion criteria, each of them will be approached in person in their respective departmental units to enroll for the study. After this, an informed written consent will be obtained by the research team before starting the study. It is anticipated that during the study, there will be attrition or addition of clinicians. Sample size has been calculated considering the attrition rate. In case of addition on clinician to units, study team will plan to give them training and mobile application using a pre-recorded training video which that can complete and obtain a certificate.

#### Blinding

Blinding to the intervention is not possible as it is a behavioural intervention.

#### Addressing confounding factors

Since there are many variations anticipated at within and between clusters, we will try to capture as many variables as possible to address the. These include patient load, bed strength, number of admissions, surgeries, details about infrastructure, mortality rate, presence of surveillance in ICU, terminal cleaning frequency, culture from clinicians at the beginning and at the end Swabs from environmental sampling, air and water, hand hygiene compliance, and compliance to standard precautions, along with detailed clinician profile.

### Sample size

Data from 653 participants would be sufficient to estimate the difference in the DDD with an SD of 13units (bnased on pilot data, unpublished), 95% confidence interval and 1 unit margin of error. Considering the clustering due to DDD being calculated from 4 sites, we consider a design effect of 1.5. Therefore, the sample size required is 1000 for the ICUs. With a slightly higher SD of 18units for DDD computed in wards and 1unit margin of error, 95% confidence interval, the sample size required is 1248. With 1.5 design effect the sample size required from wards is 1872. Data from all patients admitted during the follow-up period in the study units will be captured and included in the analysis.

### Statistical methods

Means (standard deviation), median (interquartile range), percentages will be calculated for at baseline and subsequent follow-up times. Due to the paired nature of the study, paired t-test and Chi-square tests will be used to compare baseline characteristics for continuous and categorical variables, respectively. The difference in the DDD/100 days and rates of antibiotic prescribing and variation in antibiotics dose, duration, and incidence rates of MDROs between the intervention and control units will be determined. Mixed-effects modelling will be used. The level of significance will be set at 0.05. All analysis will be done using R software. Qualitative data will be analysed using thematic framework approach.

### Outcomes

The primary outcome of the study will be calculation of defined daily dose for each antimicrobial. This will be obtained from the in-patient medical records from each unit by periodic audits during the follow-up period. Prevalence of antibiotic use among inpatients in all the study units. Our denominator will be individual units using this formula. Apart from this, the incidence of multi-drug resistance organisms (MDROs) and hospital acquired infections (HAIs) will be measured. The WHO defined dose calculator will be used as reference. The formula is given below.

1. DDD/100 beds = Number of units administered in given period (milligram) x 100 DDD (milligram WHO) x Number of days in period of study x Number of beds x occupancy index Were, occupancy index = Number of patients admitted in the ward/Total number of beds in the ward (Occupied +unoccupied)
2. For HAI Calculation of CAUTI/CLABSI/VAP (Reference: NHSN 2024) No of Central line days/Catheter days/Ventilator days DUR = No of patient days
3. For HAI Calculation of SSI (Reference: NHSN 2024) No of SSI reported SSI/100 = X 100 No of Operative procedures
4. For MDRO calculations (Reference: MDRO/CDI Module-NHSN 2024) No. Of Infections by MDRO MDRO Infection Incidence Rate = X 1000 No. Of Patient-Days

### Implications of the study

We hypothesize that an integrated m-health AMSP intervention in the form of a mobile app plus clinician training and capacity building will promote the rationale prescription of AMA and improve patient outcomes in both public and private settings. The overall goal is to strengthen the existing AMSP in the study hospitals. This is also an attempt to develop an integrated module using technology, laboratory, and hospital information to engage clinicians in AMSP. The study will also shed light on the feasibility, and sustainability issues from all the stakeholders involved in AMSP program.

## Data Availability

Not applicable as this is a protocol paper

